# Knowledge, attitudes and practices of COVID 19 among Medical Laboratory Professionals in Zambia

**DOI:** 10.1101/2020.09.22.20199240

**Authors:** Adon Chawe, Ruth L. Mfune, Paul Siyapila, Sharon D. Zimba, Pipina Vlahakis, Samson Mwale, Kapambwe Mwape, Memory Kalolekesha, Misheck Chileshe, Joseph Mutale, Tobela Mudenda, Grace Manda, Victor Daka

## Abstract

**Background:** COVID-19 is a novel disease that has spread to nearly every country worldwide. Medical laboratory professionals are key in the fight against COVID-19 as they provide confirmatory diagnosis which is the main basis upon which cases are identified and clinical management instituted. Lack of knowledge, poor attitude and unsafe laboratory practices could have negative implications towards the control of COVID-19. We conducted a quick online questionnaire to investigate the knowledge, attitudes and practices of medical laboratory personnel regarding COVID-19 in Zambia.

**Methods:** We conducted a cross sectional study among medical laboratory professionals in Zambia from 10^th^ to 29^th^ June, 2020. Data were collected using google forms and exported to SPSS version 23 for statistical analysis. Independent predictors of COVID-19 knowledge and practices were determined. Adjusted odds ratios and their 95% confidence intervals are reported.

**Results:** A total of 208 medical laboratory professionals participated in the study. There were more males (58.2%) than females. The majority of respondents had good knowledge (84.1%) and practice (75%) regarding COVID-19. Less than half (n=97, 46.6%) reported willingness to participate in a vaccination program. Predictors of good knowledge included; having a Bachelors degree (AOR: 5.0, CI: 1.15-23.9) and having prior COVID-19 related training (AOR: 8.83, CI: 2.03-38.4). Predictors of good practice included; having a masters or PhD qualification (AOR: 5.23, CI: 1.15-23.9) and having prior COVID-19 related training (AOR: 14.01, CI: 6.47-30.4).

**Conclusion:** Our findings revealed that medical laboratory professionals in Zambia have good knowledge and positive attitude towards COVID-19. However poor practices were observed There is need for continuous professional development (CPD) to ensure that medical laboratory professionals are well informed and aware of best practices to aid in curbing the pandemic.

## Introduction

Corona virus Disease also referred to as COVID-19 is a respiratory tract disease caused by SARS-CoV-2 virus [1]. Coronaviruses have been known to affect humans, infecting the respiratory tract and infections ranging from mild to severe [2]. In the past, coronaviruses have caused severe acute respiratory syndrome (SARS) and middle east respiratory syndrome (MERS), caused by SARS-CoV and MERS-CoV, respectively. Research has shown that COVID-19 is more contagious compared to the previous outbreaks, but less lethal [3]. Coughing and sneezing facilitates viral excretion and thus transmission. Inhaling these droplets or contact with infected surfaces then increases the risk of infection. There is no known vaccine or documented specific treatment for COVID 19 disease [4]. Drugs that show potential to treat critically ill patients, are still being investigated for Safety and efficacy [4–6]. Prevention and controlling the spread of COVID-19 is done by social distancing, wearing face masks to protect one from inhaling infectious droplets as well as effective hand hygiene by regularly washing hands or use of alcohol-based hand sanitizers [7,8].

The first cases of COVID-19 were reported in December 2019 in Wuhan, Hubei province, Central China. Since then, the disease has spread to nearly every country worldwide leading to the World Health Organization declaring COVID-19 a pandemic on 11^th^ March, 2020 [9]. As of 17^th^ September, 2020 there were 29 611 395 confirmed cases and 935 767 deaths reported globally [10]. In Zambia 13 887 confirmed cases with 326 deaths were reported as at 17^th^ September, 2020 [11].

Few publications and national situation reports exist that detail the number of Healthcare workers including medical laboratory personnel around the world infected with COVID-19 [12]. In China where the Disease first started 2055 healthcare workers were infected as of 24th March 2020 [13]. In the United states of America, 9282 health care workers were infected as of 14th April 2020 [14]. The World Health Organisation estimates that over 1000 health workers in Africa had been infected with COVID-19 by 23^rd^ July, 2020 [15]. Information on the source of infection among healthcare workers in countries that have recorded the disease remains limited.

Medical laboratory professionals are key personnel in the diagnosis of COVID 19 in Zambia. Although they are not in the frontline which precludes them from prominence, their role in providing confirmatory diagnosis is the main basis upon which cases are identified and clinical management instituted. The work areas of Biomedical laboratory professionals are very hazardous due to both suspected and unsuspected infectious agents. Lack of knowledge, good attitude and poor laboratory practices can have a twofold effect-a wrong diagnosis leading to wrong patient management with severe consequences or safety incidents which could be deleterious to both the concerned staff and their immediate environment which includes coworkers, families and patients or laboratory clients [16].

We conducted a study to understand the knowledge, attitudes and practices (KAP) of medical laboratory professionals in Zambia.

## Methods

### Ethical consideration

Ethical clearance for this study was obtained from the Tropical Diseases Research Centre institutional review board (IRB registration number: 00002911). The questionnaire contained an information sheet regarding the study and an informed consent statement for participants to agree to participate or not. All participants who declined to take part in the study were immediately withdrawn and could not proceed to respond to the questionnaire.

### Study design

This cross sectional survey was conducted among 208 medical laboratory professionals in Zambia. Due to COVID 19 related restrictions imposed in this period, it was not feasible to conduct face to face interviews and therefore we administered an online questionnaire using google forms. To ensure that only target respondents participated in the survey we distributed the link to the survey through the email database and WhatsApp group facilitated by the Biomedical Society of Zambia.

### Data collection

Pretesting of the questionnaire was done by administering the questionnaire to faculty at the Copperbelt University to ensure consistency and reliability of the questions and expected responses. The results from the pilot study were not included in the final analysis. The questionnaire had two main components: demographics and KAP. The demographic section had 8 questions while the KAP section was divided into the following subsections; knowledge (10 questions), attitude (5 questions) and practices (13 questions).

### Data management and statistical analysis

Collected data were downloaded and cleaned in Microsoft excel and exported to SPSS version 23 for statistical analysis. The statistical significance was set at p<0.05. To ensure the internal consistency and reliability of the data we used Cronbach alpha coefficient according to methods described [17]. To identify factors of good knowledge and practice against COVID-19, bivariate analysis was performed. All factors that were statistically significant in bivariate logistic regression were included in forward stepwise multivariate logistic regression model to identify factors that were independently associated with good knowledge and practice.

## Results

### Demographic characteristics

A total of 208 medical laboratory professionals from seven provinces of Zambia took part in this study. There were more males (n=121, 58.2%) compared to females. Half of the participants were aged 20-29 years and had a Diploma in Biomedical Science qualification. Most of the respondents were from a hospital laboratory (n=134, 64.4%) (Table 1).

**Table 1:** Demographic characteristics of participants

| Factor | Number of participants (n) | Percentage (%) |
| --- | --- | --- |
| **Sex** |  |  |
| Female | 87 | 41.8 |
| Male | 121 | 58.2 |
| **Age (years)** |  |  |
| 20 to 29 | 104 | 50.0 |
| 30 to 39 | 79 | 38.0 |
| ≥ 40 | 25 | 12.0 |
| **Current qualification** |  |  |
| Certificate in Biomedical Sciences | 2 | 1.0 |
| Diploma in Biomedical Sciences | 155 | 74.5 |
| BSc Biomedical Sciences | 40 | 19.2 |
| MSc Biomedical Sciences | 9 | 4.3 |
| PhD | 2 | 1.0 |
| **Work experience** |  |  |
| Below 5 years | 106 | 51.0 |
| 5 to 10 years | 60 | 28.8 |
| More than 10 years | 42 | 20.2 |
| **Facility type** |  |  |
| Clinic Laboratory | 14 | 6.7 |
| Health Centre Laboratory | 26 | 12.5 |
| Hospital Laboratory | 134 | 64.4 |
| Private/ NGO | 20 | 9.6 |
| Others | 14 | 6.7 |
| **Participant's province** |  |  |
| Central | 12 | 5.8 |
| Copperbelt | 61 | 29.3 |
| Eastern | 17 | 8.2 |
| Lusaka | 40 | 19.2 |
| North western | 18 | 8.7 |
| Northern | 11 | 5.3 |
| Southern | 30 | 14.4 |
| Others | 19 | 9.1 |
| **Total** | **208** | **100** |

### Factors associated to COVID-19 knowledge

A high proportion of respondents were knowledgeable on COVID-19 (n=175, 84.1%). Bivariate logistic regression analysis showed that current qualification (Crude OR:4.68, CI:1.07-20.44) and COVID-19 training (Crude OR:8.72, CI: 2.02-37.65) among participants were significantly associated with COVID-19 knowledge. Therefore, participants with higher academic qualifications and COVID-19 training were 4.68 and 8.72 times likely to have good COVID-19 knowledge respectively (Table 2).

**Table 2.** COVID-19 knowledge of participants and associated factors

| Factors | Knowledge on COVID-19 |  | Crude OR | 95% CI |
| --- | --- | --- | --- | --- |
|  | Yes (%)* | No (%)* |  |  |
| **Gender** |  |  |  |  |
| Male | 104(86.0) | 17(14.0) | 1 |  |
| Female | 71(81.6) | 16(18.4) | 0.73 | 0.34-1.53 |
| **Age (years)** |  |  |  |  |
| 20-29 | 88(84.6) | 16(15.4) | 1 |  |
| 30-39 | 65(82.3) | 14(17.7) | 0.84 | 0.39-1.85 |
| ≥ 40 | 22(88.0) | 3(12.0) | 1.33 | 0.36-4.99 |
| **Current Qualification** |  |  |  |  |
| Certificate or Diploma | 126(80.30) | 31(19.7) | 1 |  |
| Bachelors | 38(95.0) | 2(5.0) | 4.68 | 1.07-20.44 |
| Master or PhD | 11(100.0) | 0(0.0) | 316182818.76 | 0.0 |
| **Facility** |  |  |  |  |
| Clinic & health center laboratories | 32(80.0) | 8(20.0) | 1 |  |
| Hospital laboratories | 111(82.8) | 23(17.2) | 1.21 | 0.49-2.95 |
| Others | 32(94.1) | 2(5.9) | 4.00 | 0.79-20.32 |
| **Participant's province** |  |  |  |  |
| Northern | 8(72.7) | 3(27.3) | 1 |  |
| Copperbelt | 55(90.2) | 6(9.8) | 3.44 | 0.71-16.55 |
| Eastern | 13(76.5) | 4(23.5) | 1.22 | 0.22-6.92 |
| Lusaka | 32(80.0) | 8(20.0) | 1.50 | 0.32-6.97 |
| North western | 15(83.3) | 3(16.7) | 1.88 | 0.31-11.52 |
| Central | 10(83.3) | 2(16.7) | 1.88 | 0.25-14.08 |
| Southern | 27(90.0) | 3(10.0) | 3.3 | 0.57-20.10 |
| Others | 15(78.9) | 4(21.1) | 1.4 | 0.25-7.90 |
| **Trained in covid-19** |  |  |  |  |
| No | 112(78.3) | 31(21.7) | 1 |  |
| Yes | 63(96.9) | 2(3.1) | 8.72 | 2.02-37.65 |
| **Total** | **175(84.1)** | **33(15.9)** |  |  |

\* indicate that these are row percent
\*\*Others (Private laboratories, research laboratories)

### Practice towards COVID-19 by medical laboratory personnel

Poor practices towards COVID-19 were recorded among three quarters of the participants. Current qualification (Crude OR:4.51, CI:1.30-15.70), type of laboratory facility (Crude OR:3.09, CI:1.01-9.45) and COVID-19 training (Crude OR:12.97, CI:6.19-27.18) were significantly associated with good COVID-19 practices. Participants with higher academic qualifications and those with prior COVID-19 training were 4.51 and 12.97 times likely to have good COVID-19 practices (Table 3).

**Table 3.** COVID-19 practices of participants and associated factors

| Factors | Practice towards COVID-19 |  | Crude OR | 95% CI |
| --- | --- | --- | --- | --- |
|  | Poor (%)* | Good (%)* |  |  |
| **Gender** |  |  |  |  |
| Male | 92 (76.0) | 29 (24.0) | 1 |  |
| Female | 64 (73.6) | 23 (26.4) | 1.14 | 0.61-2.15 |
| **Age (years)** |  |  |  |  |
| 20-29 | 78 (75.0) | 26 (25.0) | 1 |  |
| 30-39 | 62 (78.5) | 17 (21.5) | 0.82 | 0.41-1.65 |
| ≥ 40 | 16 (64.0) | 9 (36.0) | 1.69 | 0.67-4.28 |
| **Current Qualification** |  |  |  |  |
| Certificate or Diploma | 124 (79.0) | 33 (21.0) | 1 |  |
| Bachelors | 27 (67.5) | 13 (32.5) | 1.81 | 0.84-3.89 |
| Master or PhD | 5 (45.5) | 6 (54.5) | 4.51 | 1.30-15.70 |
| **Facility** |  |  |  |  |
| Clinic & health center laboratories | 34 (85.0) | 6 (15.0) | 1 |  |
| Hospital laboratories | 100 (74.6) | 34 (25.4) | 1.93 | 0.74-4.99 |
| **Others | 22 (64.7) | 12 (35.3) | 3.09 | 1.01-9.45 |
| **Participant's province** |  |  |  |  |
| Central | 11 (91.7) | 1 (8.3) | 1 |  |
| Copperbelt | 46 (75.4) | 15 (24.6) | 3.59 | 0.43-30.14 |
| Eastern | 10 (58.8) | 7 (41.2) | 7.70 | 0.80-74.05 |
| Lusaka | 23 (57.5) | 17 (42.5) | 8.13 | 0.96-69.17 |
| North western | 16 (88.9) | 2 (11.1) | 1.38 | 0.11-17.09 |
| Northern | 10 (90.9) | 1 (9.1) | 1.10 | 0.06-20.01 |
| Southern | 25 (83.3) | 5 (16.7) | 2.20 | 0.23-21.11 |
| **Others | 15 (78.9) | 4 (21.1) | 2.93 | 0.29-30.01 |
| **Trained in COVID-19** |  |  |  |  |
| No | 129 (90.2) | 14 (9.8) | 1 |  |
| Yes | 27 (41.5) | 38 (58.5) | 12.97 | 6.19-27.18 |
| **Total** | **156 (75%)** | **52 (25%)** |  |  |

\* indicate that these are row percent
\*\*Others (Private laboratories, research laboratories)

### Attitude towards COVID-19 among medical laboratory personnel

About 93.8% reported that they would accept isolation from the community if diagnosed with COVID-19. A few (46.6%) would accept to be vaccinated against COVID-19 if a vaccine was available. On the other hand, many participants (97.6%) were ready to take part in anti-epidemic community activities. Our study revealed that 77.9% were confident that Zambia could win the battle against the COVID-19 virus (Table 4).

**Table 4.** Attitude of Biomedical professionals towards COVID-19

| Variables | Response<br>n (%) |
| --- | --- |
| Would accept isolation from the community if you were diagnosed with COVID-19] |  |
| Yes | 195 (93.8) |
| No | 8 (3.8) |
| Don't know | 5 (2.4) |
| Ready to participate in anti-epidemic in the community |  |
| Yes | 203 (97.6) |
| No | 2 (1.0) |
| Don't know | 3 (1.4) |
| Agree that COVID-19 will finally be successfully controlled |  |
| Yes | 183 (88.0) |
| No | 10 (4.8) |
| Don't know | 15 (7.2) |
| Would accept to be vaccinated against COVID-19 if a vaccine was available |  |
| Yes | 97 (46.6) |
| No | 75 (36.1) |
| I don't know | 36 (17.3) |
| Have confidence that Zambia can win the battle against the COVID-19 virus |  |
| Yes | 162 (77.9) |
| No | 24 (11.5) |
| Don't know | 22 (10.6) |
| Total | 208 (100) |

### Factors independently associated with good knowledge on COVID-19

After controlling for possible confounding, current qualification and COVID-19 training were independently associated with good COVID-19 knowledge among Biomedical professions in Zambia (Table 5).

**Table 5.** Factors independently associated with good COVID-19 knowledge

| Independent factors | Adjusted Odds Ratio | 95% CI |
| --- | --- | --- |
| ***Current Qualification*** |  |  |
| Certificate or Diploma | 1 |  |
| Bachelors | 5 | 1.13-22.19 |
| Master or PhD | 316182818.8 | 0 |
| ***Trained in covid-19*** |  |  |
| No | 1 |  |
| Yes | 8.83 | 2.03-38.44 |

### Factors independently associated with good COVID-19 practice

After controlling for possible confounding however, only current qualification and COVID-19 training were independently were associated with good COVID-19 practice among Biomedical professions in Zambia (Table 6)

**Table 6.** factors independently associated with good COVID-19 practice

| Independent factors | Adjusted Odds Ratio | 95% CI |
| --- | --- | --- |
| ***Current Qualification*** |  |  |
| Certificate or Diploma | 1 |  |
| Bachelors | 2.35 | 0.93-5.95 |
| Master or PhD | 5.23 | 1.15-23.87 |
| ***Trained in covid-19*** |  |  |
| No | 1 |  |
| Yes | 14.01 | 6.47-30.36 |

## Discussion

Very few studies worldwide have documented KAPs among HCWs towards COVID-19 due to the novel nature of the disease [18]. Our study findings showed that majority of medical laboratory professionals in Zambia had good knowledge towards COVID-19. This is comparable to findings in Vietnam and Uganda [18,19]. The majority of medical laboratory professionals had a positive attitude towards COVID-19 which is similar to findings in Vietnam [19] but contrary to other findings among healthcare workers [18,20]. The level of knowledge on COVID-19 among participants was similar irrespective of their gender, age and laboratory facility. These finding are encouraging as they indicate that laboratory professionals are knowledgeable on the transmission dynamics of the disease especially as they work with highly infectious agents. Majority of participants exhibited poor practices towards COVID-19 contrary to findings from Uganda and Nepal [18,21]. Most participants with poor practices were those who had certificate qualifications, those without prior COVID-19 training and those from clinic and health center laboratories. This could be attributed to limited resources, health information and laboratory materials in most clinic and health center laboratories found in rural areas. Poor practices can lead to delayed and/or wrong laboratory diagnosis leading to poor patient management or safety incidents that could harm the personnel and their immediate coworkers, families and patients or laboratory clients [22].

Current qualification and COVID-19 training among participants were significantly associated with good COVID-19 knowledge, a finding similar to that obtained by a study in Vietnam [19], but contrary to Bhagavathula and others [20]. Our findings show that participants with higher academic qualifications and COVID-19 training were 4.68 and 8.72 times respectively to have good COVID-19 knowledge. Current qualification was also significantly associated with good COVID-19 practices which agrees with study findings from Uganda [18]. On the other hand, type of laboratory facility and COVID-19 training were significantly associated with good COVID-19 practices. This shows that participants who had received COVID-19 training were 12.97 times likely to have good COVID-19 practices and general infection prevention. This is in agreement with a similar study in Nepal and Ethiopia [21,23].

## Limitations

The study may be susceptible to self-presentation bias as it was based on an online administered questionnaire.

## Conclusion

Our study found that medical laboratory professionals in Zambia have good knowledge and positive attitude towards COVID-19. In contrast, poor practices towards COVID-19 was found. Current qualification and COVID-19 training were independently associated with COVID-19 knowledge and practice. As the cases of COVID-19 continue to be recorded in the country, there is need for continuous professional development (CPD) among medical laboratory personnel as a key intervention in improving their contribution to COVID-19 control efforts.

## Data Availability

Data used in this manuscript is available on request.

## Acknowledgements

The authors are grateful to their families for their endurance and support during this research work.

## Authors Contributions

AC, RLM, GM and VD conceptualized the study, AC, SDZ, PV, SM, KM, MK, JM, TM developed the data collection tools. AC, PS, MC and VD performed the formal analysis and interpretation, AC, RLM, PV, KM and VD wrote the first draft manuscript. All authors read and approved the final manuscript.

## Data availability

The data analysed in this study can be made available on request

## Disclaimer

The views and opinions expressed in this article are sorely of the authors and do not reflect the official policy or position of any affiliated organization of the authors.

